# Children and Adolescent Patients with Variants in the *ATP1A3*-encoded Sodium-Potassium ATPase Alpha-3 Subunit Demonstrate an Impaired QT Response to Bradycardia and Predisposition to Sinus Node Dysfunction

**DOI:** 10.1101/2024.08.31.24312446

**Authors:** Meredith K. Srour, Minu-Tshyeto K. Bidzimou, Padmapriya Muralidharan, Saige M. Mitchell, Mary E. Moya-Mendez, Lauren E. Parker, Gabriela Reyes Valenzuela, Roberto Caraballo, Giacomo Garone, Federico Vigevano, Sarah Weckhuysen, Charissa Millevert, Monica Troncoso, Mario Matamala, Simona Balestrini, Sanjay M. Sisodiya, Josephine Poole, Claudio Zucca, Eleni Panagiotakaki, Maria T. Papadopoulou, Sébile Tchaicha, Matthildi Athina Papathanasiou Terzi, Marta Zawadzka, Maria Mazurkiewicz-Bełdzińska, Carmen Fons, Jennifer Anticona, Elisa De Grandis, Ramona Cordani, Livia Pisciotta, Sergiu Groppa, Sandra Paryjas, Francesca Ragona, Elena Mangia, Tiziana Granata, Andrey Megvinov, Rosaria Vavassori, Mohamad A. Mikati, Andrew P. Landstrom

## Abstract

**Background:** Alternating hemiplegia of childhood (AHC) is a rare disorder with both neurologic and cardiac manifestations. The ATP1A3-D801N variant is associated with a pathologically short QT interval and risk of ventricular arrhythmia following bradycardia; however, the mechanism of this remains unknown. We investigated the relationship between heart rate (HR), QT, and QTc, hypothesizing that individuals with ATP1A3-D801N have abnormal, impaired shortening of QT and QTc at lower HR leading to arrhythmia predisposition.

**Methods:** We performed a retrospective observational study of individuals who underwent clinical evaluation, Holter monitoring, and genetic testing for AHC at Duke University Hospitals. We also compiled a group of healthy individuals as a control cohort. A larger, worldwide cohort of individuals with *ATP1A3*-related phenotypes was compiled to investigate sinus node dysfunction. Linear regression analysis was then performed.

**Results:** The cohort consisted of 44 individuals with *ATP1A3*-related phenotypes with 81 Holter recordings (52.27% female; mean age at first Holter 8.04 years, range 0.58 – 33 years), compared to 36 healthy individuals with 57 Holter recordings (52.78% female; mean age at first Holter 9.84 years, range 0.08 – 38 years). Individuals with ATP1A3-D801N had reduced prolongation of QT at lower HR, manifest as a significantly lower slope for HR vs QT compared to healthy (P<0.0001). This resulted in a significantly higher slope of the relationship for HR vs QTc compared to healthy (P<0.0001). Individuals with *ATP1A3*-related phenotypes and baseline QTc <350 milliseconds (ms) had increased shortening of QT and QTc at lower HR compared to those with normal QTc (P=0.003; P=0.001). Among worldwide cases, 3 out of 131 individuals with *ATP1A3*-related phenotypes required device implantation and/or had sinus pauses >4 seconds.

**Conclusions:** Individuals with the ATP1A3-D801N variant exhibit paradoxical shortening of QT and QTc at lower HR, which contributes to an increased risk of arrhythmias during bradycardia. This is exacerbated by an underlying risk of sinus node dysfunction.

**Clinical Perspective:** What is Known:

- Individuals with ATP1A3-D801N have a short baseline QTc.
- Two individuals with AHC experienced ventricular fibrillation following bradycardia.

What the Study Adds:

- The QT and QTc shorten to a greater extent at lower heart rate in individuals with ATP1A3-D801N than in healthy individuals.
- Individuals with *ATP1A3*-related phenotypes and QTc <350ms show greater impairment of QT and QTc dynamics than those with normal QTc.
- There is low prevalence of device implantation and significant sinus pauses in individuals with *ATP1A3*-related phenotypes, with a relatively greater prevalence in those with ATP1A3-D801N.

## INTRODUCTION

Individuals with abnormally fast cardiac repolarization, manifest as a shortened QT interval, are at an increased risk of atrial and ventricular arrhythmias which can be life-threatening^1-3^. As heart rate (HR) can modulate QT interval, diagnostic criteria center around a corrected QT (QTc) which measures ventricular repolarization and normalizes it to heart rate, often using Bazett’s correction^4,5^. A diagnosis of short QT syndrome (SQTS) uses Gollob’s criteria, a point-based diagnostic system with increasing points given for QTc <370, <350, and <330ms, respectively^6^. In addition to short QTc, SQTS can also present with sinus node dysfunction, with prior case reports describing episodes of sinus arrest and junctional rhythm^7,8^. Further, there is an emerging relationship between HR and QT response in SQTS. Prior work has shown that individuals with SQTS display blunted response of QT to changes in HR when compared to healthy individuals^9-11^. However, the genetic and mechanistic causes of SQTS are often unclear. While believed to be heritable, only a minority of patients with SQTS are genotype positive for causative genes^2,12^. Thus, there is a need to identify new genes and new mechanisms of SQTS development.

Alternating hemiplegia of childhood (AHC) is a rare condition which typically manifests prior to 18 months of age with episodic hemiplegia and/or quadriplegia, dystonia, oculomotor abnormalities, seizures, dysautonomia, and sudden unexplained death in epilepsy (SUDEP)^13,14^. We have previously found that individuals with AHC have cardiac features that are similar to those with SQTS, including a shortened QTc on electrocardiogram (ECG)^15-18^ and sinus node dysfunction that may require device implantation^19,20^. Specifically, we found that individuals hosting ATP1A3-D801N, the most common missense variant among those with AHC, are uniquely predisposed to short QTc interval and ventricular arrhythmias associated with bradycardia, particularly during periods of anesthesia or sedation^15,18,21,22^. Taken together, this work suggests that SUDEP in AHC is due to the short QT phenotype; however, the link between bradycardia and arrhythmia generation remains unknown. Exploring this question will yield important new insight into SQTS, and the modulators of cardiac repolarization, more broadly.

With this in mind, we sought to investigate QT and QTc dynamics in AHC with respect to HR through linear regression analysis, which has been previously utilized for this purpose^9,23-27^. We evaluated cardiac Holter recordings of individuals with AHC and healthy individuals to determine QT and QTc responses to HR and expand our understanding of arrhythmogenic risk in ATP1A3-D801N. Finally, we convened a worldwide cohort of AHC cases to determine the risk of sinus node dysfunction and associated outcomes.

## METHODS

This was a retrospective study approved by the Duke University Hospitals Institutional Review Board, IRB #Pro00094341. Informed consent was waived. In addition, retrospective data on AHC patients was obtained using the International Consortium for Research on AHC and all the *ATP1A3* Rare Diseases (IAHCRC), according to the rules of the IAHCRC Charter. All sites had ethics board or IRB approval.

Cohorts

### Primary Cohort

The primary cohort consisted of consecutively enrolled individuals seen at Duke University Hospitals who had undergone clinical evaluation and genetic testing for AHC. Individuals were included if they 1) met full Aicardi diagnostic criteria for AHC^13^, 2) met atypical/partial AHC diagnostic criteria and, 3) were diagnosed with cerebellar ataxia, areflexia, pes cavus, optic atrophy, and sensorineural deafness (CAPOS), another disorder associated with *ATP1A3* mutations^17^. Individuals were excluded if they did not have 1) a reported ECG, or 2) an ambulatory Holter monitoring report using HScribe^TM^ software. Individuals in this cohort were further categorized based on genotype as follows: ATP1A3-D801N; other pathogenic or likely pathogenic single nucleotide variant in *ATP1A3*, defined as ATP1A3 non-D801N; presumed loss of function variant in *ATP1A3* such as deletion or splice site variant, defined as *ATP1A3* LOF; and no likely pathogenic or pathogenic *ATP1A3* variant detected on genetic testing, defined as genotype negative or *ATP1A3* Neg.

### Healthy Cohort

Healthy individuals were selected for a comparison cohort if they were seen at Duke University Hospitals for a cardiovascular evaluation. The following individuals were excluded: 1) elite athletes with documented cardiovascular changes secondary to intense cardiovascular activity; 2) those with known congenital heart disease, channelopathy, cardiomyopathy, and skeletal myopathy; 3) those with known genetic syndrome with cardiac phenotype; 4) those taking cardiac medications, including antiarrhythmics; 5) those with an abnormal ECG and/or echocardiogram; and 6) those with ventricular or supraventricular ectopy >1% of total QRS complexes on Holter recording. All healthy individuals had undergone Holter monitoring with HScribe^TM^ software following cardiology visits for risk prediction due to identification of a genetic variant associated with cardiac disease, family history of sudden cardiac arrest, cardiomyopathy, and/or channelopathy, or history of arrhythmias or prolonged QT that had since resolved. Healthy individuals were matched to AHC individuals based on sex and age (+/- 2 years).

### IAHCRC Cohort

Given the rarity of AHC diagnoses, a multinational rare disease registry was established which consisted of individuals from Duke University Hospitals as above, the IAHCRC, and the Baylor College of Medicine (BCM). The cohort from the IAHCRC has been previously described^28^. Inclusion criteria for these individuals included, 1) meeting full Aicardi diagnostic criteria for AHC as diagnosed by a neurologist^13^, 2) meeting atypical/partial AHC diagnostic criteria, and 3) meeting criteria for other diagnoses, such as rapid-onset dystonia-parkinsonism (RDP) or CAPOS, if they had a confirmed variant in *ATP1A3*^28^. Individuals were excluded from analysis if they 1) did not meet the inclusion criteria, 2) did not have outcome data available, such as survival and pacemaker (PM) or PM/implantable cardioverter defibrillator (ICD) implantation, and 3) did not have at least 1 ECG with original tracings available for re-review.

### Holter Data Analysis

Holter data was collected from the Primary Cohort and Healthy Cohort using the HScribe^TM^ cardiac monitoring software. Holter recordings ranged in duration from 45 minutes to 24 hours. Automatically generated minimum, mean, and maximum HR, QT, and QTc measurements over 5-minute intervals throughout the Holter recording were abstracted. QTc was calculated using Bazett’s correction^5^. A linear regression analysis was performed for each Holter recording and genotype, and a line of fit equation was calculated. Automated QT or QTc measurements that appeared to be outliers on visual inspection of linear regression and were greater than 1.5 times the value of the line of fit equation for a given HR were remeasured manually using Lepeschkin’s method and Bazett’s correction^5,29^ and were excluded if they differed. To validate the automated Holter recording measurements, >15% of total Holter recordings were manually re-evaluated by two raters blinded to genotype using Lepeschkin’s method and Bazett’s correction^5,29^. The average QT and QTc for every HR interval for each genotype were then assessed for interrater agreement, defined as a difference of ≤ 10% of the lower measurement, and Bland-Altman analysis was performed. If there was >10% difference between measurements, the average of both measurements was used for analysis. As a second method of validation, all ECGs for individuals in the cohort were included in a replication analysis. The HR, QT, and QTc for each ECG were manually measured by an electrophysiologist blinded to genotype. Linear regression analyses of HR vs QT and QTc for individuals grouped by genotype were then performed.

### Outcomes Analysis

We used Kaplan-Meier analysis to assess freedom from composite events, defined as 1) requiring a PM or PM/ICD, or 2) having documented sinus pauses greater than 4 seconds, based on age at last follow-up. For the Primary Cohort, this age was defined as age at last cardiology visit. For the IAHCRC Cohort, last follow-up was defined as November 1, 2022, as the database was last updated through October 2022. For those who had a composite event, age was defined as when they required device implantation or first had sinus pauses greater than 4 seconds. For Holter analyses, we only included individuals with Holter data available for review.

### Statistical Analysis

Statistical analysis was performed using both JMP Pro 17 and Prism 10. Results are reported as mean ± standard deviation with 95% confidence intervals unless otherwise noted. A Kruskal-Wallis test with Dunn’s post-hoc multiple comparisons or Dunnet’s post-hoc comparison to healthy was used for continuous variables with more than two groups. Fisher’s exact test was used for nominal variables with more than two groups. Mann-Whitney test was used for continuous variables with two groups. Wilcoxon matched pairs signed rank test was used to compare two groups with paired samples. Simple linear regression analysis was modeled by the equation QT or QTc = mHR + b, where m is equivalent to slope and b is equivalent to intercept. This analysis was conducted for overall linear regression for each genotype, as well as individual linear regressions for each Holter recording and first diagnostic Holter recording. A mixed model was also used to verify the genotype linear regression and account for repeated measurements from the same individual. The HR, genotype, and interaction between HR and genotype were fixed effects, QT and QTc were outcome variables, and subject number was a random effect with a residual repeated structure. Kaplan-Meier analysis was compared using log-rank test. Significance was set at P= 0.05.

## RESULTS

### Primary Cohort Characteristics

To determine the relationship between cardiac repolarization time and HR, we first evaluated individuals with AHC and healthy individuals recruited from Duke University Hospitals. In total, 44 individuals with *ATP1A3*-related phenotypes and 36 healthy individuals were included in the Primary Cohort (**Table 1**). All Holter recordings available in the HScribe^TM^ software were utilized. Individuals with *ATP1A3*-related phenotypes were grouped based on genotype as follows: ATP1A3-D801N (9 individuals, 25 Holter recordings); ATP1A3 non-D801N (25 individuals, 38 Holter recordings); *ATP1A3* LOF (4 individuals, 9 Holter recordings); *ATP1A3* Neg (6 individuals, 9 Holter recordings). In the healthy group, 36 individuals with 57 Holter recordings were included. We found no significant difference in mean age at first diagnostic Holter and biological sex between genotypes (**Table 1**). Overall, <1% of data points from Holter recordings were excluded.

**Table 1.**
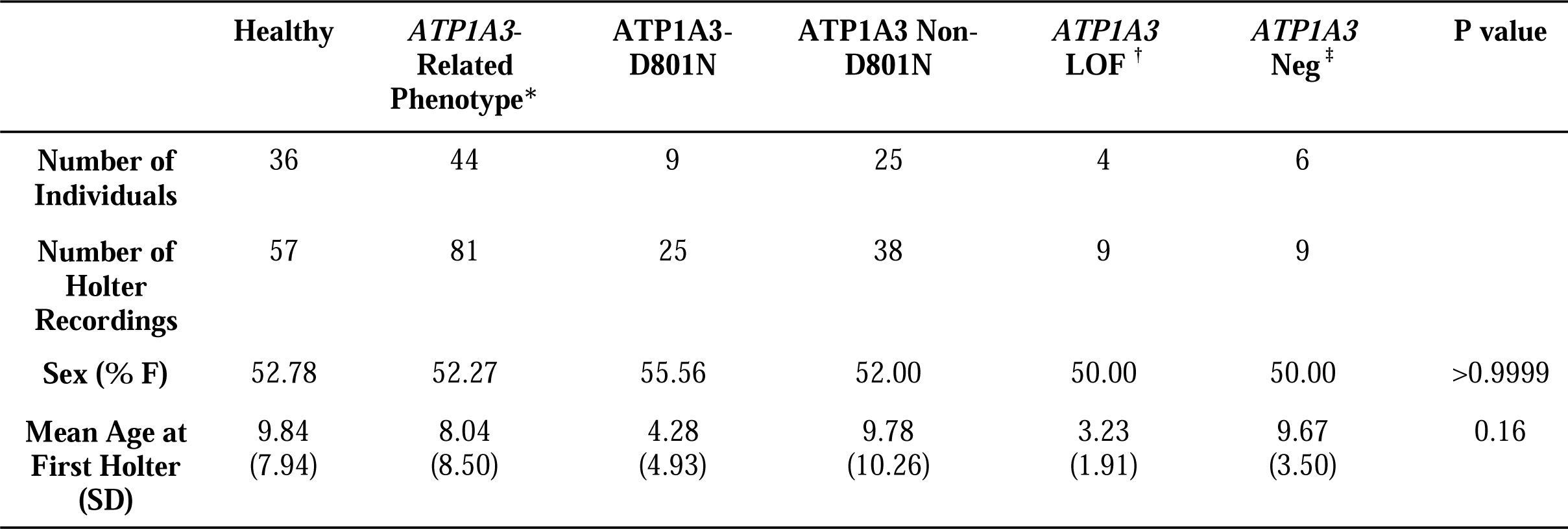
Primary cohort characteristics. *Includes individuals with a formal diagnosis of AHC (n=37) or CAPOS (n=1), those who did not meet full criteria for AHC but were found to host an *ATP1A3* variant (n=5), and one individual with no variant in *ATP1A3* who did not meet full criteria for AHC (n=1). †Includes individuals with deletion (n=3) or splice site variant in *ATP1A3* (n=1). ‡Includes individuals with no pathogenic or likely pathogenic variant in *ATP1A3* identified (n=6). P values were calculated using Fisher’s exact test or Kruskal-Wallis test. LOF, loss of function variant; Neg, genotype negative.

### Individuals with ATP1A3-D801N show blunted QT change to heart rate variability

Our group has previously shown that individuals with ATP1A3-D801N have shorter QTc on ECG compared to those with other variants^15^. Similarly, in our cohort, individuals with ATP1A3-D801N had an overall mean QT and QTc on Holter that were significantly lower than that of healthy individuals (302.68 + 19.27 and 385.60 + 27.95, vs 341.12 + 32.36 and 426.96 + 20.54, respectively; P <0.0001). Further, the mean QTc for ATP1A3-D801N was significantly lower than that of *ATP1A3* LOF and *ATP1A3* Neg. We then investigated HR modulation of QT interval in each group using a linear regression analysis. It is well established that the QT interval increases with reduced HR and decreases with increased HR^4,30^ and that the QTc, which is corrected for HR, remains relatively constant across a range of HRs in healthy individuals^4,5^. We confirmed this observation in our healthy cohort which showed prolonged QT with slower HR and normalization of HR with QTc, demonstrated by a mean slope of HR vs QT of -1.52 and a mean slope of HR vs QTc of 0.22 (**Figures 1 and 2**; **Table 2**). Conversely, we found that individuals with ATP1A3-D801N exhibited diminished QT prolongation at lower HRs (**Figure 1; Figure S1; Table S1**). Specifically, we found that D801N probands had a lower mean linear regression slope than healthy individuals (-0.85 ± 0.46 vs -1.52 ± 0.42, respectively; P <0.0001; **Table 2**). For example, based on our model, at a mean HR of 60 beats per minute (bpm), D801N probands had a QT of 335.33ms. Conversely, healthy individuals had a mean QT of 396.64ms (**Table S2**). These results were unique to individuals hosting the D801N variant. For example, individuals in the ATP1A3 non-D801N, *ATP1A3* LOF, and *ATP1A3* Neg genetic subgroups showed a similar QT vs HR relationship to healthy individuals (mean slope -1.24 ± 0.43, -1.41 ± 0.35, -1.41 ± 0.16, respectively, vs -1.52 ± 0.42; P value not significant). Further, each of these genotype groups showed less impairment of QT dynamics compared to ATP1A3-D801N (**Table 2**). To account for different numbers of Holter recordings per individual, this analysis was also replicated using only the first diagnostic Holter and demonstrated a similar relationship (**Figure S2; Table S3**). We then validated the automated QT measurements using manual measurements with 2 independent evaluators blinded to genotype. Again, we observed a blunted response of QT to lower HR in ATP1A3-D801N with strong interrater agreement (see **Supplemental Results; Figures S3-S4, Table S4**). An independent validation using manual HR and QT/QTc measurements from clinical ECGs blinded to genotype demonstrated a similar relationship (**Figure S5**).

**Figure 1.** ATP1A3-D801N is associated with blunted response of QT to lower HR compared to healthy. Linear regression analysis of mean HR versus mean QT over five-minute intervals throughout each Holter recording for individuals in the primary and healthy cohorts. Colored points represent individual data points for all Holter recordings included in each genotype group. Solid line represents overall linear regression line of fit. m represents slope of the line of fit equation. A. Healthy cohort (n=57), QT= -1.64HR + 501.40; R^2^= 0.70. B. ATP1A3-D801N subgroup (n=25), QT= -0.79HR + 381.50; R^2^= 0.32. C. ATP1A3 non-D801N subgroup (n=38), QT= -1.27HR + 445.10; R^2^= 0.45. D. *ATP1A3* LOF subgroup (n=9), QT= -1.28HR + 465.50; R^2^= 0.61. E. *ATP1A3* Neg subgroup (n=9), QT= -1.44HR + 483.80; R^2^= 0.80. F. Overall lines of fit for each of the respective subgroups. Blue, healthy; black, ATP1A3-D801N; red, ATP1A3 non-D801N; purple, *ATP1A3* LOF; orange, *ATP1A3* Neg.

**Figure 2.** ATP1A3-D801N is associated with increased shortening of QTc at lower HR compared to healthy. Linear regression analysis of mean heart rate versus mean QTc over five-minute intervals throughout Holter recording for individuals in the primary and healthy cohorts. Colored points represent individual data points for all Holter recordings included in each genotype group. Solid line represents overall linear regression line of fit. m represents slope of the line of fit equation. A. Healthy (n=57), QTc= 0.06HR + 421.80; R^2^= 0.0023. B. ATP1A3-D801N (n=25), QTc= 0.96HR + 291.40; R^2^= 0.30. C. ATP1A3 non-D801N (n=38), QTc= 0.42HR + 362.10; R^2^= 0.055. D. ATP1A3 LOF (n=9), QTc= 0.41HR + 390.40; R^2^= 0.083. E. ATP1A3 Neg (n=9), QTc= 0.28HR + 406.30; R^2^= 0.093. F. Lines of fit for all groups. Blue, healthy; black, ATP1A3-D801N; red, ATP1A3 non-D801N; purple, *ATP1A3* LOF; orange, *ATP1A3* Neg.

**Table 2.** Mean slope of linear regression for HR versus QT and QTc, compared by genotype. *Healthy compared to ATP1A3-D801N, using Kruskal-Wallis test with Dunnet’s post-hoc comparison to healthy. Values listed as mean + SD (95% confidence interval). HR, heart rate; QTc, corrected QT; LOF, loss of function variant; Neg, genotype negative.

|  | **Healthy** | **ATP1A3-D801N** | **ATP1A3 Non-D801N** | **ATP1A3 LOF** | **ATP1A3 Neg** | **P value** |
| --- | --- | --- | --- | --- | --- | --- |
| **n** | 57 | 25 | 38 | 9 | 9 |  |
| **Mean Slope**<br>**HR vs QT** | -1.52 ± 0.42<br>(-1.63 – -1.41) | -0.85 ± 0.46<br>(-1.03 – -0.67) | -1.24 ± 0.43<br>(-1.37 – -1.10) | -1.41 ± 0.35<br>(-1.64 – -1.19) | -1.41 ± 0.16<br>(-1.52 – -1.30) | <0.0001* |
| **Mean Slope**<br>**HR vs QTc** | 0.22 ± 0.43<br>(0.11 – 0.33) | 0.80 ± 0.51<br>(0.60 – 1.00) | 0.52 ± 0.46<br>(0.38 – 0.67) | 0.20 ± 0.50 (-<br>0.13 – 0.53) | 0.32 ± 0.22<br>(0.18 – 0.47) | <0.0001* |

\*Healthy compared to ATP1A3-D801N, using Kruskal-Wallis test with Dunnet's post-hoc comparison to healthy. Values listed as mean ± SD (95% confidence interval). HR, heart rate; QTc, corrected QT; LOF, loss of function variant; Neg, genotype negative.

As several variables, including biological sex, can influence cardiac repolarization time^5,31^, we next conducted a sub-analysis to determine whether other variables might influence this finding. We found that females with an *ATP1A3*-related phenotype, and those with ATP1A3 non-D801N variants, had a significantly lower slope for HR vs QT compared to males, although it was a mild physiologic difference. Further, there were no significant differences between male and females in all other groups (**Table S5**). When analyzed by age, there was no significant difference in QT dynamics between individuals younger than 10, 10–20, or older than 20 years old for ATP1A3-D801N or all *ATP1A3*-related phenotypes (**Table S6**). Taken together, these findings suggest that individuals with ATP1A3-D801N have a blunted prolongation of QT at lower HRs, while other genetic subtypes of AHC were more similar to healthy individuals in their QT dynamics.

### Individuals with ATP1A3-D801N show paradoxical shortening of QTc interval at lower heart rates

Given the diminished QT shortening with lower HRs demonstrated in the ATP1A3-D801N group, we hypothesized that this group has an accentuated shortening of QTc at low HR when compared to healthy individuals. Indeed, while control individuals demonstrated no significant change in QTc at lower heart rates, we found that ATP1A3-D801N individuals had further shortening of the QTc (**Figure 2; Figure S6; Table S1**), demonstrated by a greater mean slope than healthy individuals (0.80 ± 0.51 vs 0.22 ± 0.43, respectively; P <0.0001; **Table 2**). For example, at a heart rate of 60bpm, the QTc for ATP1A3-D801N was 356.7ms compared to 415.6ms in healthy individuals (**Table S7**). As with QT, this relationship was similar when replicated using only the first diagnostic Holter recordings for each individual and upon manual measurement validation (**Supplemental Results; Figures S7-S9, Tables S3 and S8**). When linear regression was performed for ECGs, there was also increased shortening of QTc at lower HR for ATP1A3-D801N variant-positive individuals (**Figure S10).** Further, this blunted response was seen equally in both D801N-positive males and females (**Table S9**), and there was no difference in QTc dynamics based on age in individuals with *ATP1A3*-related phenotypes (**Table S10**). In summary, these results suggest that the blunted QT prolongation with slower HR among individuals with ATP1A3-D801N is associated with a paradoxical reduction of QTc at lower HRs compared to healthy controls and all other genetic subtypes of *ATP1A3*-related phenotypes. We conclude that, given the already low QTc at baseline in these individuals, bradycardia causes further reduction in QTc to a markedly abnormal range, exacerbating a known arrhythmic risk factor.

### Individuals with QTc<350ms demonstrate blunted QT prolongation and QTc shortening with bradycardia

We then investigated whether the maladaptive QT and QTc response to low HR is an ATP1A3-D801N specific phenomenon, or whether individuals with other *ATP1A3* variants who have short QTc at baseline also have this abnormal response. To do this, we separated individuals with *ATP1A3*-related phenotypes into different groups defined by their baseline QTc on diagnostic ECG. Groups were defined using Gollob’s criteria^6^, where normal QTc was between 370-470ms, borderline short QTc was between 350-370ms, and short QTc was <350ms. Seven of the 44 patients with *ATP1A3*-related phenotypes had QTc <350ms. Four of these hosted ATP1A3-D801N and the other 3 hosted ATP1A3 non-D801N variants (**Table S11**). None of the individuals with *ATP1A3* LOF, *ATP1A3* Neg, or healthy individuals had QTc <350ms (**Table S11**). We found that those with short diagnostic QTc (<350ms) had a diminished QT prolongation response to HR compared to those with normal QTc (370-470ms), and the slope of HR vs QTc in the short QTc group was significantly greater than that of the normal QTc group (**Figure 3; Table S11**). When separated by specific genotype, there was greater impairment of HR vs QT dynamics in ATP1A3-D801N than in ATP1A3 non-D801N for those with QTc <350ms. However, within each genotype, there was no significant difference in QT or QTc dynamics when grouped by baseline QTc (see **Supplemental Results; Tables S12-S13**). Taken together, we found that presence of short QTc of <350ms on diagnostic ECG for all *ATP1A3*-related phenotypes is associated with greater impairment of QT and QTc dynamics. While this effect was independent of genotype, most patients with QTc <350 hosted the ATP1A3-D801N variant.

**Figure 3.** Short baseline QTc is associated with greater impairment of QT and QTc dynamics. Each colored line represents the linear regression for Holter recordings from individuals in the primary and healthy cohorts with a given baseline QTc on ECG. A. Mean HR versus mean QT for Holter recordings from individuals with *ATP1A3*-related phenotypes (n=81) compared to healthy patients (n=57). B. Mean HR versus mean QTc for Holter recordings from individuals with *ATP1A3*-related phenotypes (n=81) compared to healthy (n=57). Black, <350ms (n=16); red, 350-370ms (n=15); purple, 370-470ms (n=50); blue, healthy, all 370-470ms (n=57).

### Individuals with *ATP1A3* variants are predisposed to significant sinus node dysfunction

Given the association between bradycardia and QTc shortening, we next determined whether patients hosting *ATP1A3* variants were at increased risk of sinus node dysfunction^19^. To do this, we leveraged a large, multinational registry of individuals with AHC and *ATP1A3* genetic variants. The cohort was comprised of 131 individuals who met our inclusion/exclusion criteria: the same 44 individuals from Duke University Medical Center, 86 individuals from the IAHCRC, and 1 individual from BCM previously described in the literature^15^. Of these additional 87 individuals, 31 had available Holter recording measurements. Additional demographic characteristics for individuals from the IAHCRC and BCM are detailed in **Table S14**. We found no difference in sex or age representation among the genetic subgroups (**Table S14**). Individuals with ATP1A3-D801N reached an average maximum HR during Holter monitoring that was significantly lower than healthy individuals (153.94 ± 18.80 vs 176.21 ± 22.52; P <0.0001). Otherwise, there was no significant difference in average minimum HR, mean HR, and ventricular and supraventricular ectopy burden between any of the groups and healthy individuals (**Table 3**). We identified 3 out of 131 individuals who met the composite endpoint of implantation of a PM or PM/ICD or documented sinus pause, with a mean age at follow-up of 15.62 years (maximum of 50 years). Among them, 2 out of 3 hosted the D801N variant and the third hosted ATP1A3-S137F. For healthy individuals, *ATP1A3* LOF, and *ATP1A3* Neg, no individuals met the composite endpoint during the follow-up period. This yielded a calculated composite event rate of 0.15%/year/individual for the entire cohort, with a D801N-positive event rate of 0.38%/year/individual (**Figure 4**). Thus, leveraging the largest international cohort to date to explore this endpoint, we concluded that, although the frequency of significant sinus node dysfunction is low, individuals hosting the D801N variant have a greater predisposition to developing it compared to other genetic subtypes.

**Figure 4.** ATP1A3-D801N is associated with sinus node dysfunction. A. Kaplan-Meier analysis of freedom from composite events versus age of event in individuals from primary and IAHCRC cohorts (n=131), as compared to those from the healthy cohort (n=36). Tick marks represent censored individuals. Bars represent 95% confidence intervals. No significant difference between healthy and *ATP1A3*-related phenotype using log-rank test (P = 0.57). B. Characteristics of individuals who had composite events. PM, pacemaker; ICD, implantable cardioverter defibrillator.

**Table 3.** Holter recording characteristics for all individuals from primary and IAHCRC cohorts. Heart rates are listed in beats per minute (BPM), RR is listed in seconds. *Listed as median (interquartile range), as one individual had 1102 VE per hour (23157 total). P value calculated with Fisher’s exact test or Kruskal-Wallis test with Dunn’s post-hoc multiple comparisons. HR, heart rate; VE, ventricular ectopy; SVE, supraventricular ectopy; LOF, loss of function variant; Neg, genotype negative.

|  | **Healthy** | **ATP1A3-D801N** | **ATP1A3 Non-D801N** | **ATP1A3 LOF** | **ATP1A3 Neg** | **P value** |
| --- | --- | --- | --- | --- | --- | --- |
| **Number of Individuals (Holter recordings)** | 36<br>(57) | 19<br>(36) | 47<br>(58) | 5<br>(10) | 7<br>(10) |  |
| **Sex (% Female)** | 52.78 | 57.89 | 53.19 | 60.00 | 57.14 | 0.98 |
| **Mean Age at First Holter (SD)** | 9.84<br>(7.94) | 13.75<br>(12.58) | 10.98<br>(10.16) | 5.35<br>(5.02) | 12.85<br>(9.00) | 0.44 |
| **Minimum HR (SD)** | 57.18<br>(8.30) | 51.39<br>(7.48) | 55.72<br>(9.84) | 55.80<br>(8.92) | 59.60<br>(5.38) | 0.03 |
| **Mean HR (SD)** | 95.61<br>(6.53) | 92.54<br>(12.98) | 93.95<br>(16.80) | 104.60<br>(10.12) | 94.90<br>(8.74) | 0.18 |
| **Maximum HR (SD)** | 176.21<br>(22.52) | 153.94<br>(18.80) | 159.05<br>(19.84) | 166.70<br>(17.11) | 168.10<br>(16.67) | **<0.0001** |
| **Longest RR (SD)** | 1.26<br>(0.22) | 1.57<br>(0.48) | 1.30<br>(0.24) | 1.48<br>(0.61) | 1.28<br>(0.21) | **0.008** |
| **VE/Hour** | 0.50<br>(3.29) | 0.00*<br>(0.08) | 0.51<br>(3.55) | 0.37<br>(1.10) | 0.00<br>(0.00) | 0.11 |
| **SVE/Hour** | 0.12<br>(0.24) | 1.65<br>(7.25) | 2.62<br>(14.38) | 0.22<br>(0.71) | 0.034<br>(0.048) | 0.09 |

### Flunarizine may exacerbate the blunted QT response to bradycardia but does not worsen sinus node dysfunction

Given that the standard of care for treatment of AHC involves the use of flunarizine^32,33^, a T-type calcium channel blocker, we next explored whether use of flunarizine was associated with exacerbation of the cardiac phenotype observed, since its impact on the heart is unknown. Of the 131 individuals included, 90 were taking flunarizine and there was no significant difference between flunarizine use and the genetic subtypes (**Table S15**). Among them, flunarizine use was not associated with changes in average minimum, mean, or maximum HR for any genotype group when compared to those who were not taking flunarizine (**Table S15**). When exploring the relationship between QT/QTc dynamics and flunarizine, we did find that the overall slope for HR vs QTc was significantly greater for individuals with all *ATP1A3*-related phenotypes taking flunarizine than for those who were not (**Table S15**). Specifically, we found that at a HR of 60 bpm, the mean QTc was 376.97ms among those taking flunarizine versus 402.4ms among those not taking flunarizine. This suggests that flunarizine might be associated with impairment of QT prolongation during periods of bradycardia; however, the physiologic and clinical relevance of this finding is uncertain. When separated by biological sex, the mean slope for HR vs QT among individuals taking flunarizine was significantly lower in females than in males. At 60bpm, the mean QT in females taking flunarizine was 350.9ms versus 366.1ms in males, suggesting that flunarizine shortening of the QT interval might be exacerbated in female patients (**Table S16**). This sex-bias was also observed for all patients with *ATP1A3*-related phenotypes, regardless of flunarizine use (**Tables S5 and S9**). There was no difference in QT/QTc dynamics based on flunarizine use for males (**Table S17**). However, females taking flunarizine had a significantly lower slope for HR vs QT than females who were not (**Table S18**). Taken together, flunarizine may statistically impair QT dynamics during lower HR among specific subgroups of patients, particularly among female individuals. Despite this statistical difference, the overall physiologic impact of flunarizine on the heart remains unclear. This is particularly important as there is a clear role for flunarizine in treating the neurologic manifestations of *ATP1A3*-related disease.

## DISCUSSION

This study expands our understanding of arrhythmia risk in individuals with *ATP1A3*-related diseases, such as AHC. In healthy individuals, the QT interval adapts to changes in HR by shortening when the HR increases and lengthening when the HR decreases^4,30^. When corrected by HR, the QTc remains relatively fixed across a range of HRs among healthy individuals. We demonstrate that individuals with the ATP1A3-D801N variant show impairment of this relationship with decreased prolongation of QT at lower HR, similar to individuals diagnosed with SQTS^9,10^. This phenomenon has previously been described as deceleration-dependent QT interval shortening^34,35^. One mechanistic explanation for this finding is that increased acetylcholine inhibits the proper prolongation of QT when the HR slows^34,36^. Additionally, inhibition of autonomic activity has been shown to cause impairment of QT dynamics^37^. Dysautonomia is a minor diagnostic criterion for AHC^13^ and could therefore be an explanation for the findings we describe in ATP1A3-D801N. Overall, we uncover a potential novel mechanism of QT modulation that could increase arrhythmia risk at lower HR, which seems to be based in a myocardial defect. However, the specific details of how autonomic alterations further exacerbate risk is still unknown.

Correcting QT for HR can be done through a variety of different means^5,38-40^. Previous work has demonstrated limitations to Bazett’s correction in effectively normalizing the QTc during bradycardia and tachycardia^41,42^. Importantly, we found that Bazett’s correction held for all healthy individual HR ranges, which include most HR ranges in our disease cohorts. Specifically, the slope of mean HR vs mean QTc for healthy individuals was close to zero, indicating effective normalization of the QT for differences in HR (**Figure 2**). From this, we ultimately concluded that the automated Bazett’s correction formula was suitable for correcting QT in all groups. In addition, we assessed association of changes in QT and QTc dynamics with factors such as biological sex, age, and flunarizine use. While we found a difference in HR vs QT between males and females, there was no difference in HR vs QTc, suggesting that HR variation might manifest as a difference in QT responsiveness that subsides with QT correction for HR. Age did not seem to have an effect, whereas flunarizine use showed a small effect on HR vs QTc dynamics. However, we see no clear evidence that flunarizine negatively impacts the heart. Moreover, it is the medication of choice for alleviation of neurologic symptoms in AHC, therefore the known benefits are significantly greater than any harms.

Given the finding that individuals with ATP1A3-D801N have short QTc at baseline^15,16^, which is a known arrhythmia risk^1-3^, we sought to determine the effect of heart rate on this QT abnormality. We found that individuals with *ATP1A3*-related phenotypes who also had a short baseline QTc demonstrated impairment of normal QT dynamics with changing HR, particularly during periods of bradycardia. This results in a shortening of the QTc at lower HR, which exacerbates the short QT phenotype further. It is likely that this is the proarrhythmic trigger for many patients who have had a life-threatening arrhythmic event with *ATP1A3*-related disease. As such, it is critical that patients with AHC, and anyone with an *ATP1A3* mutation, are evaluated by an electrophysiology and cardiovascular genetic specialist to determine their QT/QTc at baseline. This clinical evaluation, when combined with genetic risk assessment, is key to determining the risk for arrhythmias. Furthermore, with increasing knowledge of the cardiac manifestations of AHC, questions arise as to whether these individuals are at risk of sinus node dysfunction requiring PM or ICD implantation. Analysis of our combined cohort shows that sinus node dysfunction is uncommon, with few individuals requiring a device or experiencing significant sinus pauses. While the risk is low, events can be severe when they do occur, and our results indicate a greater event rate in those with ATP1A3-D801N. Thus, it is crucial to universally check and routinely monitor sinus node dysfunction in these individuals through ambulatory monitoring. Further, as sinus bradycardia resulting from sinus node dysfunction might compound the risk in patients with the D801N variant and short QT, this may represent another pro-arrhythmic variable.

We acknowledge limitations to our study design and methodology. As this is a retrospective study, it is unclear if abnormal QT and QTc dynamics with decreasing HR are directly responsible for arrhythmias in individuals with ATP1A3-D801N. Future directions include exploring rate dependency in human induced pluripotent stem cell cardiomyocytes (iPSC-CMs) from individuals with ATP1A3-D801N to better characterize this mechanism. Additionally, AHC is a rare disease and our cohort is small. As a result, we may be underpowered to detect certain differences. However, this is the largest study to date of Holter recordings from patients with AHC across a multinational registry. Finally, rather than using beat-to-beat data, our study utilizes five-minute averages of HR, QT, and QTc provided by the Holter monitoring software. Although this provides us with less data than would otherwise be available, we were still able to detect a significant difference in relationships between ATP1A3-D801N and healthy.

## CONCLUSIONS

Overall, our findings support the idea that AHC, and specifically the ATP1A3-D801N variant, can manifest as a cardiac phenotype resembling SQTS. It is important to advise individuals with ATP1A3-D801N, particularly those with very short baseline QTc on ECG, of the risk of arrhythmia with slowed HR, such as during sedation or anesthesia. Currently, there is no treatment for abnormal QT and QTc dynamics, although quinidine has been proposed for patients with SQTS^10^. For now, cardiac monitoring during periods of possible low HR is advised.

## Non-Standard Abbreviations and Acronyms

AHC: alternating hemiplegia of childhood
*ATP1A3*: gene encoding for the alpha-3 subunit of sodium-potassium ATPase
ATP1A3-D801N: D801N missense variant in *ATP1A3*-encoded ATP1A3
ATP1A3 Non-D801N: missense single nucleotide variants, not including D801N, in *ATP1A3*-encoded ATP1A3
*ATP1A3* LOF: presumed loss of function variants in *ATP1A3*-encoded ATP1A
*ATP1A3* Neg: no pathogenic or likely pathogenic variant identified in *ATP1A3*
QTc: corrected QT interval
HR: heart rate
BPM: beats per minute
SUDEP: sudden unexplained death in epilepsy

## Data Availability

Dr. Andrew Landstrom had full access to all the data in the study and takes responsibility for data integrity and accuracy of data analysis.

## ACKNOWLEDGEMENTS

Thank you to all contributors to the IAHCRC. Dr. Andrew Landstrom had full access to all the data in the study and takes responsibility for data integrity and accuracy of data analysis.

## SUPPLEMENTAL MATERIAL

Tables S1–S18

Figures S1-S10

