## Supplemental Materials for "Children and Adolescent Patients with Variants in the *ATP1A3*-encoded Sodium-Potassium ATPase Alpha-3 Subunit Demonstrate an Impaired QT Response to Bradycardia and Predisposition to Sinus Node Dysfunction"

**SUPPLEMENTAL MATERIAL**

**SUPPLEMENTAL RESULTS**

**Validation of automated measurements**

To ensure accuracy in QT and QTc measurements, we validated greater than 10% of the automated Holter recordings using manual measurements. The manually validated cohort consisted of 23 individuals with 25 total Holter recordings: 7 from the ATP1A3-D801N group (8 Holter recordings), 6 from the ATP1A3 non-D801N group (7 Holter recordings), 2 from the *ATP1A3* LOF group (2 Holter recordings), 2 from the *ATP1A3* Neg group (2 Holter recordings), and 6 from the healthy group (6 Holter recordings). Two individuals (MS and SM) conducted blinded, manual re-measurements. The overall relationship between HR and QT/QTc for ATP1A3-D801N was similar using manual measurements, with increased shortening of the QT and QTc at lower heart rates (**Figures S3 & S8**). There was no significant difference in the mean slope for HR vs QTc between automated and manual measurements, with acceptable interrater agreement as demonstrated by the majority of measurements falling within 95% limits of agreement (**Figures S4 & S9; Tables S4 & S8**).

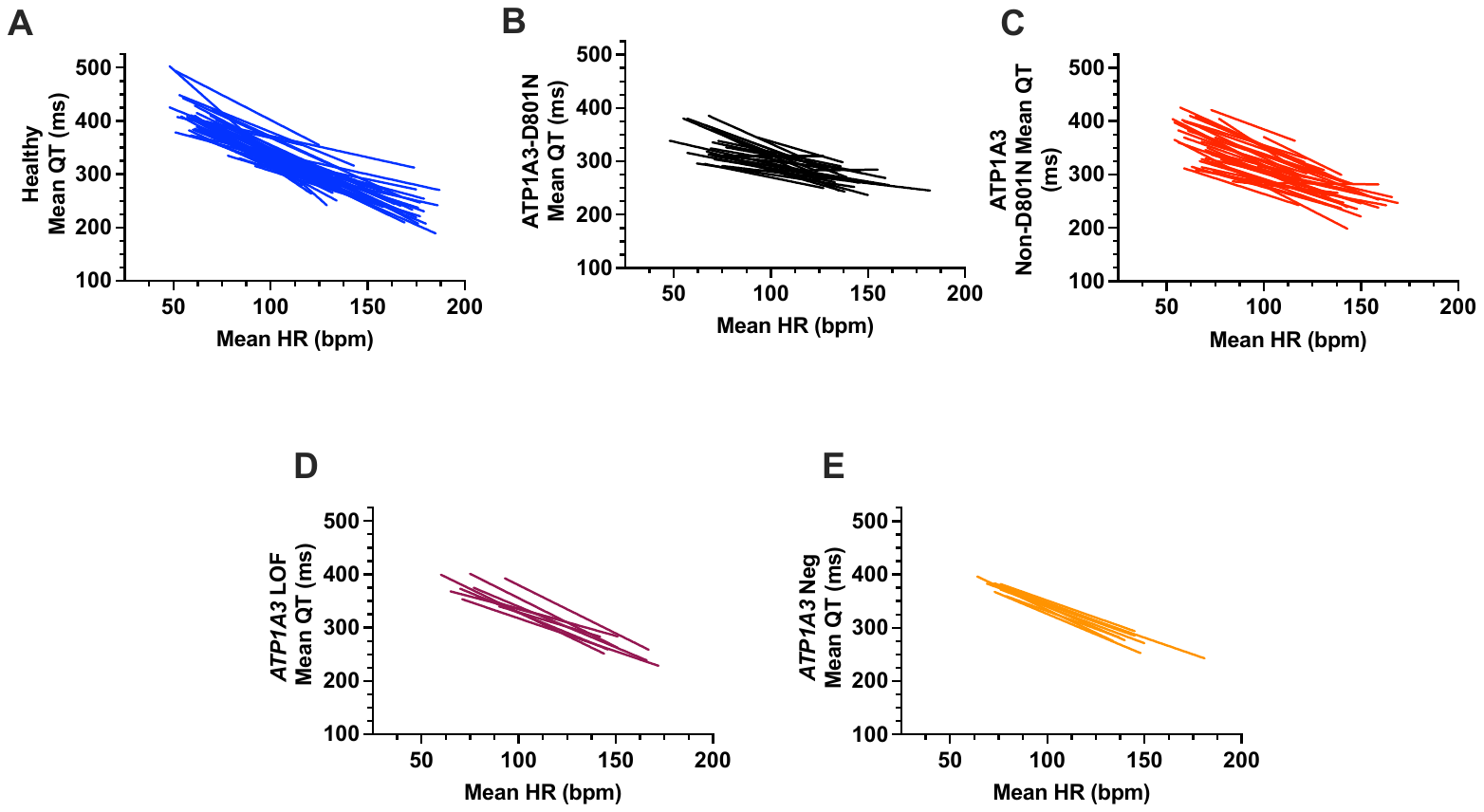
**SUPPLEMENTAL FIGURES**

**Figure S1. ATP1A3-D801N is associated with impaired prolongation of QT at lower heart rate compared to healthy.** Graphs of QT intervals by heart rate (HR). Lines represent linear regression lines of fit for individual Holter recordings from individuals within each subgroup. ATP1A3-D801N showed impaired prolongation of QT at lower HR compared to healthy. **A.** Healthy (n=57). **B.** ATP1A3-D801N (n=25). **C.** ATP1A3 Non-D801N (n=38). **D.** *ATP1A3* LOF (n=9). **E.** *ATP1A3* Neg (n=9). Blue, healthy; black, ATP1A3-D801N; red, ATP1A3 non-D801N; purple, *ATP1A3* LOF; orange, *ATP1A3* Neg.

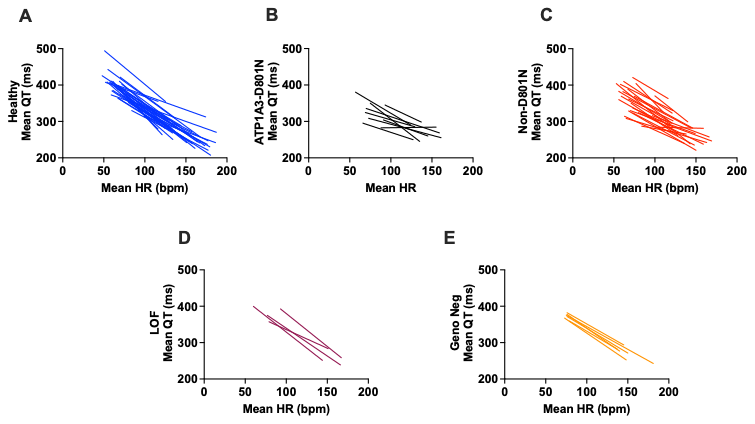

**Figure S2. First diagnostic Holter recordings demonstrate that ATP1A3-D801N is associated with blunted QT prolongation at lower heart rates compared to healthy controls and other *ATP1A3* genotypes.** Graphs of QT interval measurements by heart rate (HR). Lines represent linear regression lines of fit for first diagnostic Holter recordings. Again, there is reduced prolongation of the QT interval in ATP1A3-D801N at lower HR. **A.** Healthy (n=36). **B.** ATP1A3-D801N (n=9). **C.** ATP1A3 Non-D801N (n=25). **D.** *ATP1A3* LOF (n=4). **E.** *ATP1A3* Neg (n=6). Blue, healthy; black, ATP1A3-D801N; red, ATP1A3 non-D801N; purple, *ATP1A3* LOF; orange, *ATP1A3* Neg.

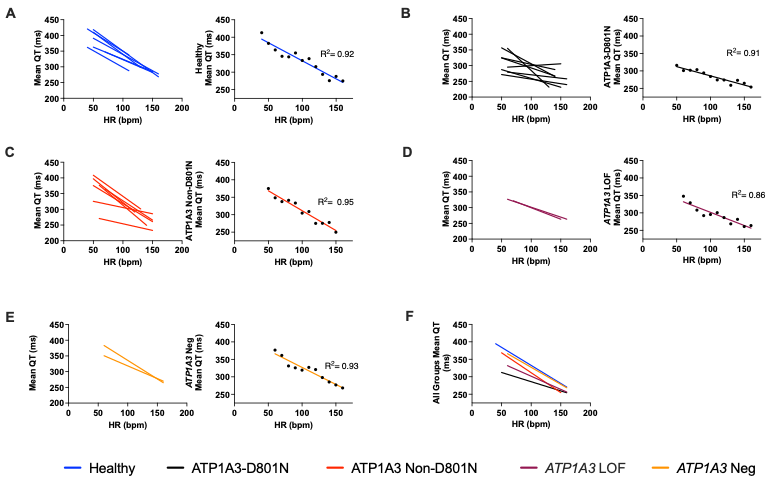

**Figure S3. Manual measurement validation demonstrates reduced QT prolongation at lower heart rates in ATP1A3-D801N.** Left panel shows linear regression lines of fit for individual Holter recordings, comprised of manually measured QT measurements. Right panel shows overall fit line for all Holter recordings from all patients in the respective genotype group, with R^2^ listed. Each black point represents a heart rate (HR) and mean QT pair validated between two raters. Colors correspond to each genotype. Similar to the automated findings, ATP1A3-D801N shows impaired prolongation of QT at lower HR. **A.** Healthy (n=6). **B.** ATP1A3-D801N (n=8). **C.** ATP1A3 Non-D801N (n=7). **D.** *ATP1A3* LOF (n=2). **E.** *ATP1A3* Neg (n=2). **F.** All groups. Blue, healthy; black, ATP1A3-D801N; red, ATP1A3 non-D801N; purple, *ATP1A3* LOF; orange, *ATP1A3* Neg.

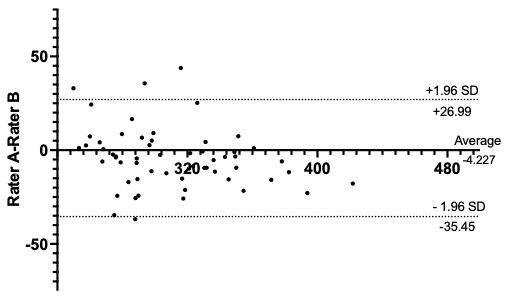
**Figure S4. Bland-Altman analysis shows acceptable interrater agreement for manual measurements of heart rate versus mean QT.** There is a bias of -4.23, standard deviation of bias of 15.93, and 95% limits of agreement from -35.45 to 26.99. Y-axis represents difference in QT measurements between Rater A and Rater B. X-axis represents average of QT measurements of Rater A and Rater B.

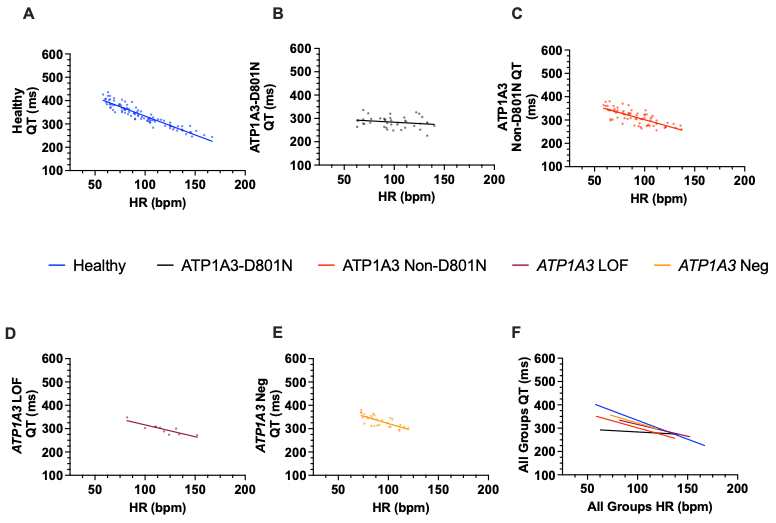

**Figure S5. Manually measured QT interval on ECG demonstrates blunted increase at lower heart rates in ATP1A3-D801N.** Graphs of manually measured QT intervals on ECG by heart rate (HR), grouped by genotype. Solid lines represent overall linear regression lines of fit. Points represent measurements for individuals in the cohort. A. Healthy (n=35, 93 ECGs). B. ATP1A3-D801N (n=9, 40 ECGs). C. ATP1A3 non-D801N (n=25, 74 ECGs). D. *ATP1A3* LOF (n=4, 10 ECGs). E. *ATP1A3* Neg (n=6, 28 ECGs). F. Overall lines of fit for each of the respective subgroups. Blue, healthy; black, ATP1A3-D801N; red, ATP1A3 non-D801N; purple, *ATP1A3* LOF; orange, *ATP1A3* Neg.

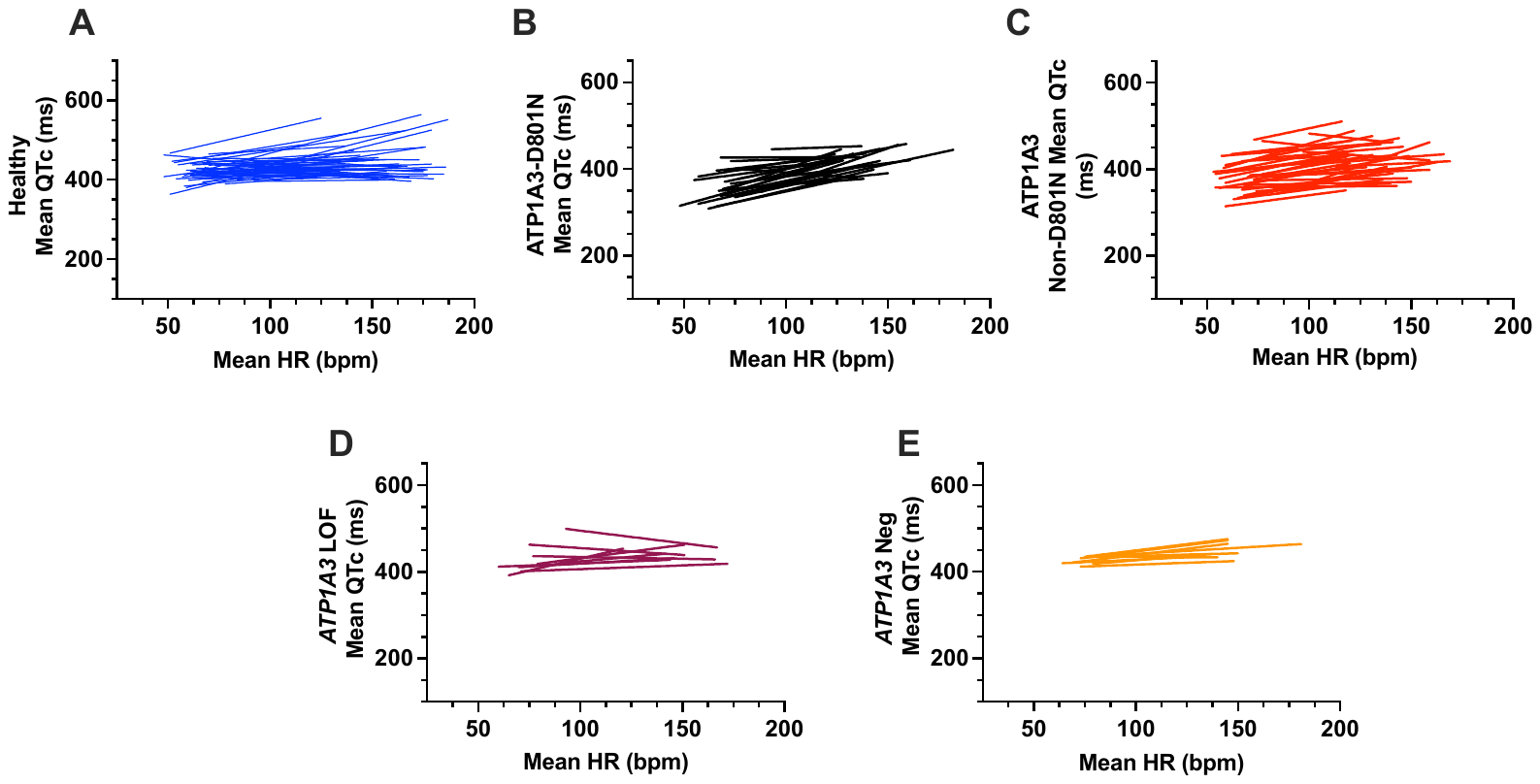

**Figure S6. ATP1A3-D801N is associated with paradoxical shortening of QTc at lower heart rate compared to healthy.** Graphs of QTc intervals by heart rate (HR). Lines represent linear regression lines of fit for individual Holter recordings from individuals within each subgroup. ATP1A3-D801N showed increased shortening of QTc at slower HR compared to healthy. **A.** Healthy (n=57). **B.** ATP1A3-D801N (n=25). **C.** ATP1A3 Non-D801N (n=38). **D.** *ATP1A3* LOF (n=9). **E.** *ATP1A3* Neg (n=9). Blue, healthy; black, ATP1A3-D801N; red, ATP1A3 non-D801N; purple, *ATP1A3* LOF; orange, *ATP1A3* Neg.

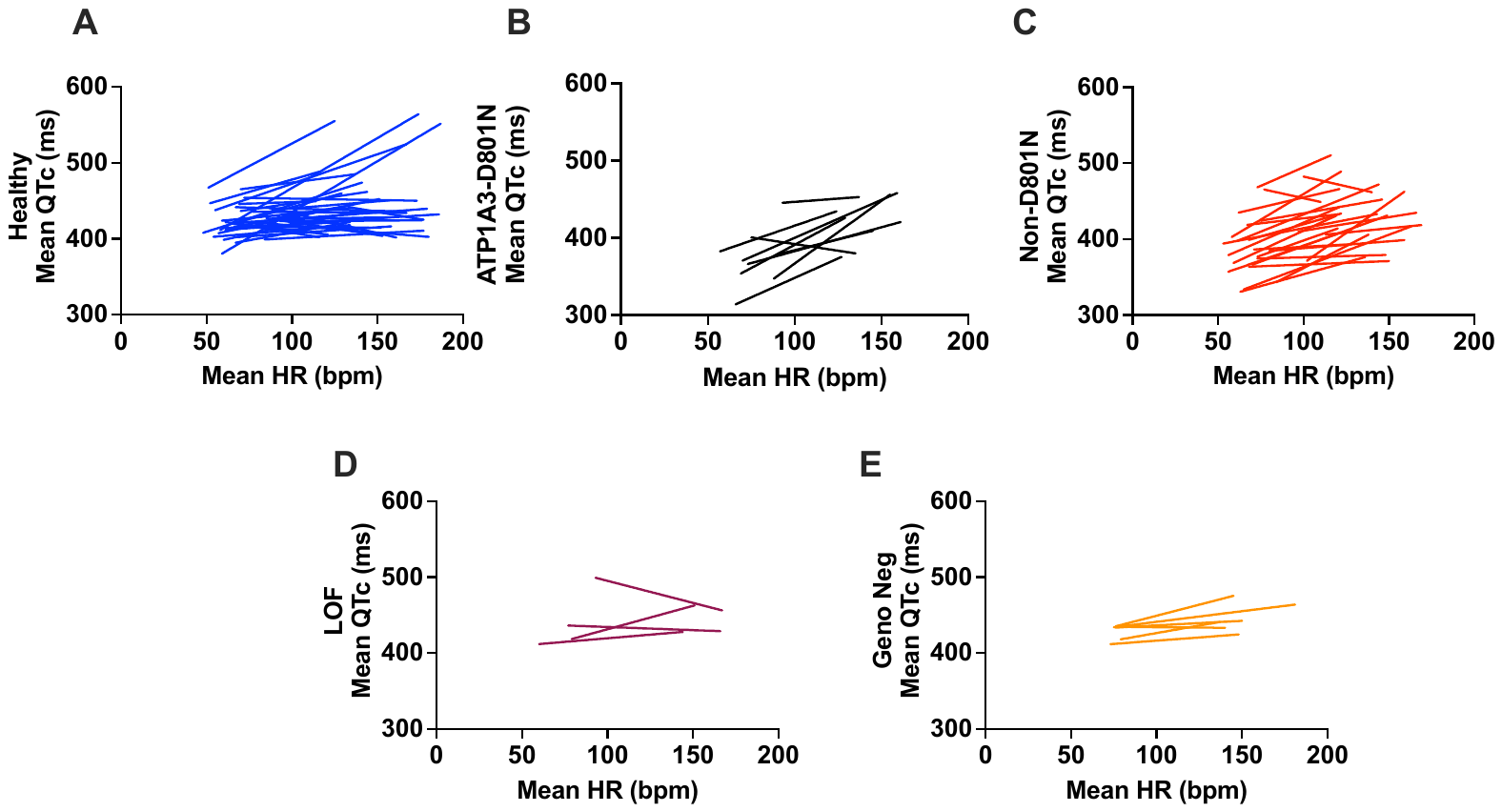

**Figure S7. First diagnostic Holter recordings similarly indicate that ATP1A3-D801N is associated with increased shortening of QTc at lower heart rate compared to healthy.**

Graphs of QTc intervals by heart rate (HR). Lines represent linear regression lines of fit for first diagnostic Holter recordings. The first diagnostic Holter recordings show a similar relationship to all recordings included. **A.** Healthy (n=36). **B.** ATP1A3-D801N (n=9). **A.** Healthy (n=36). **B.** ATP1A3-D801N (n=9). **C.** ATP1A3 Non-D801N (n=25). **D.** *ATP1A3* LOF (n=4). **E.** *ATP1A3* Neg (n=6). Blue, healthy; black, ATP1A3-D801N; red, ATP1A3 non-D801N; purple, *ATP1A3* LOF; orange= *ATP1A3* Neg.

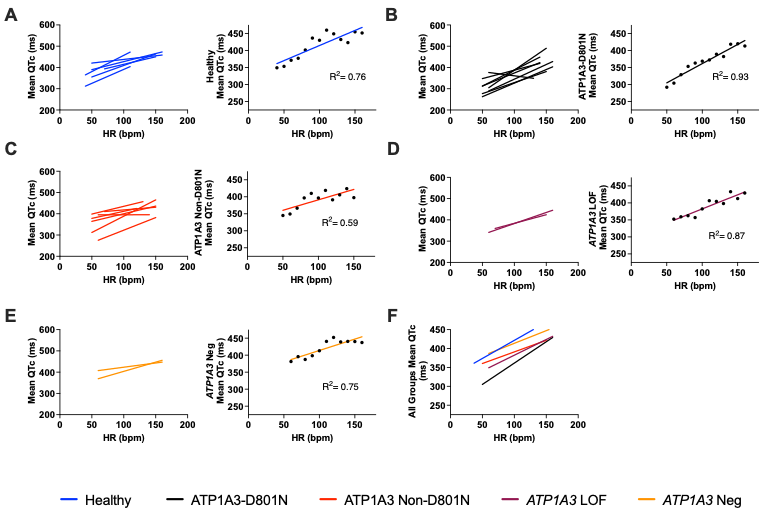

**Figure S8. Manual measurement validation shows increased shortening of QTc at lower heart rates in ATP1A3-D801N.** Left panel shows linear regression lines of fit for individual Holter recordings, comprised of manually measured QTc measurements by one rater. Right panel shows overall line of fit for all Holter recordings from all patients in that genotype group, with R^2^ listed. Each black point represents a heart rate (HR) and mean QTc pair validated between two raters. Colors correspond to each genotype. **A.** Healthy (n=6). **B.** ATP1A3-D801N (n=8). **C.** ATP1A3 Non-D801N (n=7). **D.** *ATP1A3* LOF (n=2). **E.** *ATP1A3* Neg (n=2). **F.** All groups.

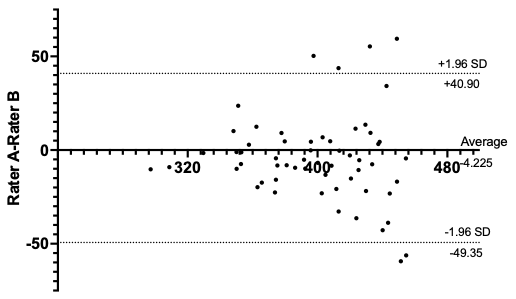

**Figure S9. Bland-Altman analysis shows acceptable interrater agreement for manual measurements of heart rate and mean QTc.** There is a bias of -4.23, standard deviation of bias of 23.02 and 95% limits of agreement from -49.35 to 40.90. Y-axis represents difference in QTc measurements between Rater A and Rater B. X-axis represents average of QTc measurements of Rater A and Rater B.

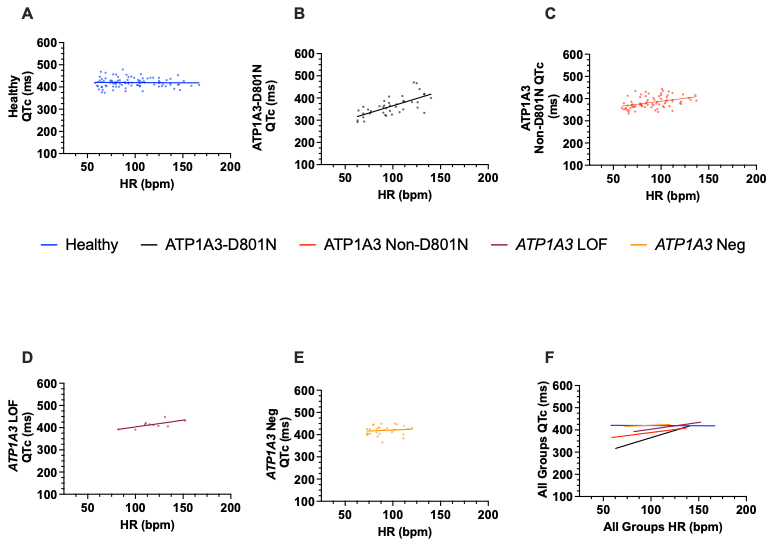

**Figure S10. Manual measurement of QTc on ECGs demonstrates paradoxical shortening at lower heart rates in ATP1A3-D801N.** Graphs of manual QTc intervals on ECG by heart rate (HR), grouped by genotype. Solid lines represent overall linear regression lines of fit. Points represent measurements for individuals in the cohort. A. Healthy cohort (n=35, 93 ECGs). B. ATP1A3-D801N (n=9, 40 ECGs). C. ATP1A3 non-D801N (n=25, 74 ECGs). D. *ATP1A3* LOF (n=4, 10 ECGs). E. *ATP1A3* Neg (n=6, 28 ECGs). F. Overall lines of fit for each of the respective subgroups. Blue, healthy; black, ATP1A3-D801N; red, ATP1A3 non-D801N; purple, *ATP1A3* LOF; orange, *ATP1A3* Neg.

**SUPPLEMENTAL TABLES**

**Table S1. Slope of overall line of fit for simple linear regression compared to mixed model for each genotype.**

|  | **Healthy**  **(n=57)** | **ATP1A3-D801N**  **(n=25)** | **ATP1A3 Non-D801N**  **(n=38)** | ***ATP1A3* LOF**  **(n=9)** | ***ATP1A3* Neg**  **(n=9)** |
| --- | --- | --- | --- | --- | --- |
| **Slope HR vs QT**  **Linear Regression** | -1.65 | -0.79 | -1.27 | -1.28 | -1.44 |
| **Slope HR vs QT**  **Mixed Model** | -1.52 | -0.82 | -1.17 | -1.32 | -1.39 |
| **Slope HR vs QTc**  **Linear Regression** | 0.05 | 0.96 | 0.42 | 0.41 | 0.28 |
| **Slope HR vs QTc**  **Mixed Model** | 0.21 | 0.89 | 0.53 | 0.33 | 0.33 |

HR, heart rate; QTc, corrected QT; LOF, loss of function variant; Neg, genotype negative.

**Table S2. QT values for heart rates of 60 and 100 beats per minute by genotype, based on mean linear regression equations for all Holter recordings in each group.**

| **Genotype** | **Equation** | **QT at HR 60 bpm (ms)** | **QT at HR 100 bpm (ms)** |
| --- | --- | --- | --- |
| **Healthy** | QT= -1.52(HR) + 487.77 | 396.64 | 335.89 |
| **ATP1A3-D801N** | QT= -0.85(HR) + 386.30 | 335.33 | 301.34 |
| **ATP1A3 Non-D801N** | QT= -1.24(HR) + 439.10 | 364.92 | 315.46 |
| ***ATP1A3* LOF** | QT= -1.41(HR)+ 480.37 | 395.48 | 338.89 |
| ***ATP1A3* Neg** | QT= -1.41(HR) + 480.50 | 395.98 | 339.63 |

Equations are shown where slope is equivalent to mean slope and y-intercept is equivalent to mean y-intercept for the individual linear regression equations of all Holter recordings included in the analysis. HR, heart rate; bpm, beats per minute; ms, milliseconds; LOF, loss of function; Neg, genotype negative.

|  | **Healthy** | **ATP1A3-D801N** | **ATP1A3 Non-D801N** | ***ATP1A3* LOF** | ***ATP1A3* Neg** | **P value** |
| --- | --- | --- | --- | --- | --- | --- |
| **n** | 36 | 9 | 25 | 4 | 6 |  |
| **Mean Slope**  **HR vs QT** | -1.49 $\pm$ 0.39  (-1.62 –  -1.36) | -0.85 $\pm$ 0.50  (-1.18 –  -0.53) | -1.20 $\pm$0.48  (-1.38 –  -1.01) | -1.53 $\pm$ 0.36  (-1.88 –  -1.18) | -1.39 $\pm$ 0.13  (-1.49 –  -1.28) | **0.001*** |
| **Mean Slope**  **HR vs QTc** | 0.24 $\pm$ 0.44  (0.10 – 0.39) | 0.73 $\pm$ 0.58  (0.35 – 1.11) | 0.57 $\pm$ 0.52 (0.36 – 0.77) | 0.04 $\pm$ 0.50  (-0.46 – 0.53) | 0.25 $\pm$ 0.21 (0.08 – 0.43) | **0.03*** |

**Table S3. Mean slope of linear regression for HR versus QT and QTc for first diagnostic Holter recordings grouped by genotype.**

Values listed as mean $\pm$ SD (95% confidence interval). *: Healthy compared to ATP1A3-D801N, Kruskal-Wallis test with Dunnet’s post-hoc comparison to healthy. HR, heart rate; QTc, corrected QT; LOF, loss of function variant; Neg, genotype negative.

|  | **Automated** | **Manual** | **P value** |
| --- | --- | --- | --- |
| **No. Holter recordings** | 25 | 25 |  |
| **Mean slope** | -1.08 $\pm$ 0.52  (-1.28 – -0.87) | -0.88 $\pm$0.47  (-1.06 – -0.69) | **0.01** |

**Table S4. Validation of automated and manual mean linear regression slope for HR versus QT.**

Values listed as mean $\pm$ SD (95% confidence interval), P value calculated with Wilcoxon matched-pairs signed rank test.

**Table S5. Mean slope of linear regression for HR versus QT for first diagnostic Holter recording, grouped by genotype and biological sex.**

|  | **Healthy** | ***ATP1A3*-Related Phenotype** | **ATP1A3 D801N** | **ATP1A3 Non-D801N** | ***ATP1A3* LOF** | ***ATP1A3***  **Neg** |
| --- | --- | --- | --- | --- | --- | --- |
| **n** | 36 | 44 | 9 | 25 | 4 | 6 |
| **Male** | -1.48 $\pm$0.44  (-1.69 – -1.27) n=17 | -1.30 $\pm$ 0.44  (-1.49 – -1.11) n=21 | -0.67 $\pm$0.56  (-1.22 – -0.12) n=4 | -1.41 $\pm$0.27  (-1.56 – 1.26) n=12 | -1.65 $\pm$ 0.17  (-1.88 – -1.41) n=2 | -1.48 $\pm$ 0.13  (-1.63 – -1.33) n=3 |
| **Female** | -1.49 $\pm$ 0.35  (-1.64 – -1.33) n=19 | -1.08 $\pm$0.49  (-1.28 – -0.88) n=23 | -1.00 $\pm$ 0.44  (-1.39 – -0.61) n=5 | -1.00 $\pm$0.55  (-1.30 – -0.70) n=13 | -1.42 $\pm$ 0.56  (-2.19 – -0.64) n=2 | -1.33 $\pm$ 0.04  (-1.37 – -1.28) n=3 |
| **P value** | 0.70 | **0.03** | 0.41 | **0.007** | >0.9999 | 0.40 |

Values listed as mean $\pm$ SD (95% confidence interval), P values calculated with Mann-Whitney test. HR, heart rate; LOF, loss of function variant; Neg, genotype negative.

| **Age (years)** | **Healthy** | ***ATP1A3*-Related Phenotypes** | **ATP1A3-D801N** | **All Non-D801N Genotypes** |
| --- | --- | --- | --- | --- |
| **<10** | -1.44 $\pm$ 0.29  (-1.54 – -1.34)  n= 32 | -1.07 $\pm$ 0.47  (-1.20 – -0.95)  n=55 | -0.80 $\pm$ 0.49  (-1.03 – -0.58)  n=18 | -1.20 $\pm$ 0.40  (-1.33 – -1.07)  n=37 |
| **10 – 20** | -1.64 $\pm$ 0.52  (-1.86 – -1.42) n=22 | -1.30 $\pm$ 0.36  (-1.44 – -1.15)  n=22 | -0.97 $\pm$ 0.36  (-1.23 – -0.70)  n=7 | -1.45 $\pm$ 0.24  (-1.57 – -1.33)  n=15 |
| **>20** | -1.48 $\pm$ 0.75  (-2.33 – -0.63)  n=3 | -1.55 $\pm$ 0.56  (-2.10 – -1.00)  n=4 | -- | -1.55 $\pm$ 0.56  (-2.10 – -1.00)  n=4 |
| **P value** | 0.29 | 0.05 | 0.39 | **0.048** |

**Table S6. Comparison of mean slope for HR versus QT based on age.**

Values listed as mean $\pm$ SD (95% confidence interval). P values calculated using Kruskal-Wallis with Dunn’s multiple comparisons or Mann-Whitney test. HR, heart rate.

**Table S7. QTc values at heart rates of 60 and 100 beats per minute by genotype, based on mean linear regression equations for all Holter recordings in each group.**

| **Genotype** | **Equation** | **QTc at HR 60 bpm (ms)** | **QTc at HR 100 bpm (ms)** |
| --- | --- | --- | --- |
| **Healthy** | QTc= 0.22(HR) + 402.22 | 415.57 | 424.46 |
| **ATP1A3-D801N** | QTc= 0.80(HR) + 308.82 | 356.68 | 388.59 |
| **ATP1A3 Non-D801N** | QTc= 0.52(HR) + 354.15 | 385.60 | 406.57 |
| ***ATP1A3* LOF** | QTc= 0.20(HR) + 415.20 | 427.14 | 435.10 |
| ***ATP1A3* Neg** | QTc= 0.32(HR) + 402.99 | 422.37 | 435.28 |

Equations are calculated where slope is equivalent to mean slope and y-intercept is equivalent to mean y-intercept for the individual linear regression equations of all Holter recordings included in the analysis. HR, heart rate; QTc, corrected QT; bpm, beats per minute; ms, milliseconds; LOF, loss of function; Neg, genotype negative.

|  | **Automated** | **Manual** | **P value** |
| --- | --- | --- | --- |
| **No. Holter recordings** | 25 | 25 |  |
| **Mean slope** | 0.73 $\pm$ 0.50  (0.53– 0.92) | 0.91 $\pm$ 0.54  (0.70 – 1.12) | 0.06 |

**Table S8. Validation of automated and manual mean linear regression slope for HR versus QTc.**

Values listed as mean $\pm$ SD (95% confidence interval), *P* value calculated with Wilcoxon matched-pairs signed rank test. HR, heart rate.

**Table S9. Mean slope of linear regression for HR versus QTc for first diagnostic Holter recording, grouped by genotype and biological sex.**

Values listed as mean $\pm$ SD (95% confidence interval), P values calculated with Mann-Whitney test. HR, heart rate; QTc, corrected QT; LOF, loss of function variant; Neg, genotype negative

|  | **Healthy** | ***ATP1A3*-Related Phenotype** | **ATP1A3 D801N** | **ATP1A3 Non-D801N** | ***ATP1A3* LOF** | ***ATP1A3***  **Neg** |
| --- | --- | --- | --- | --- | --- | --- |
| **n** | 36 | 44 | 9 | 25 | 4 | 6 |
| **Male** | 0.15 $\pm$ 0.44  (-0.06 – 0.36) n=17 | 0.44 $\pm$ 0.48 (0.23 – 0.65) n=21 | 1.00 $\pm$ 0.44 (0.57 – 1.43) n=4 | 0.38 $\pm$ 0.44 (0.13 – 0.63) n=12 | 0.05 $\pm$ 0.19  (-0.21 – 0.32) n=2 | 0.18 $\pm$ 0.21  (-0.05 – 0.42) n=3 |
| **Female** | 0.32 $\pm$0.44 (0.12 – 0.51) n=19 | 0.59 $\pm$ 0.56 (0.36 – 0.81) n=23 | 0.52 $\pm$ 0.63  (-0.03 – 1.07) n=5 | 0.74 $\pm$ 0.54 (0.44 – 1.03) n=13 | 0.02 $\pm$ 0.85  (-1.15 – 1.19) n=2 | 0.43 $\pm$ 0.27 (0.13 – 0.73) n=3 |
| **P value** | 0.43 | 0.22 | 0.41 | 0.05 | >0.9999 | 0.40 |

| **Age (years)** | **Healthy** | ***ATP1A3*-Related Phenotypes** | **ATP1A3-D801N** | **All Non-D801N Genotypes** |
| --- | --- | --- | --- | --- |
| **<10** | 0.07 $\pm$0.26  (-0.02 – 0.16) n=32 | 0.51 $\pm$ 0.52  (0.38 – 0.65)  n=55 | 0.76 $\pm$ 0.57  (0.50 – 1.02)  n=18 | 0.39 $\pm$ 0.45  (0.25 – 0.54)  n=37 |
| **10 – 20** | 0.42 $\pm$ 0.50  (0.21 – 0.62)  n=22 | 0.61 $\pm$ 0.38  (0.45 – 0.77)  n=22 | 0.89 $\pm$ 1.14  (0.65 – 1.14)  n=7 | 0.48 $\pm$ 0.34  (0.31 – 0.66)  n=15 |
| **>20** | 0.43 $\pm$ 0.84  (-0.52 – 1.38)  n=3 | 0.70 $\pm$ 0.82  (-0.10 – 1.50)  n=4 | -- | 0.70 $\pm$ 0.82  (-0.10 – 1.50)  n=4 |
| **P value** | **0.006*** | 0.46 | 0.47 | 0.30 |

**Table S10. Comparison of mean slope for HR versus QTc based on age.**

Values listed as mean $\pm$ SD (95% confidence interval). P values calculated using Kruskal-Wallis with Dunn’s multiple comparisons or Mann-Whitney test. *: significant difference between <10 and 10–20, P =0.005. HR, heart rate; QTc = corrected QT.

| **Baseline QTc (ms)** | **<350ms** | **350-370ms** | **370-470ms** | **P value** |
| --- | --- | --- | --- | --- |
| **Healthy** | 0 (0) | 0 (0) | 36 (57) | -- |
| **ATP1A3-D801N** | 4 (10) | 5 (7) | 4 (8) | -- |
| **ATP1A3 Non-D801N** | 3 (6) | 6 (8) | 18 (24) | -- |
| ***ATP1A3* LOF** | 0 (0) | 0 (0) | 4 (9) | -- |
| ***ATP1A3* Neg** | 0 (0) | 0 (0) | 6 (9) | -- |
| **Healthy HR vs QT** | -- | -- | -1.52 $\pm$0.42  (-1.63 – -1.41) | **0.006*** |
| ***ATP1A3-*Related Phenotype**  **HR vs QT** | -0.87$\pm$ 0.43  (-1.08 – -0.66) | -1.05$\pm$ 0.47  (-1.69 – -0.81) | -1.28 $\pm$ 0.42  (-1.39 – -1.16) | **0.003**^†^ |
| **Healthy**  **HR vs QTc** | -- | -- | 0.22 $\pm$0.43  (0.11 – 0.33) | **0.01*** |
| ***ATP1A3-*Related Phenotype**  **HR vs QTc** | 0.89 $\pm$ 0.38  (0.70 – 1.08) | 0.64 $\pm$ 0.47  (0.40 ­– 0.88) | 0.42 $\pm$ 0.48  (0.28 – 0.55) | **0.001**^†^ |

**Table S11. Mean linear regression slope for HR versus QT and QTc, grouped by baseline QTc on ECG in *ATP1A3-*related phenotypes and healthy.**

Values listed as individuals (Holter recordings) or mean $\pm$ SD (95% confidence interval)

*: compared to *ATP1A3-*related phenotypes, Mann-Whitney test. †: <350ms vs 370-470ms, Kruskal-Wallis test with Dunn’s post-hoc multiple comparisons. HR, heart rate; QTc, corrected QT; LOF, loss of function variant; Neg, genotype negative.

**Table S12. Mean linear regression slope for HR versus QT grouped by baseline QTc on ECG.**

| **Baseline QTc (ms)** | **Healthy** | **ATP1A3-D801N** | **ATP1A3**  **Non-D801N** | ***ATP1A3* LOF** | ***ATP1A3* Neg** | **P value** |
| --- | --- | --- | --- | --- | --- | --- |
| **<350** | -- | -0.65 $\pm$ 0.31  (-0.84 – -0.46) | -1.23 $\pm$ 0.36  (-1.67 – -0.80) | -- | -- | **0.003** |
| **350-370** | -- | -0.77 $\pm$0.53  (-1.16 ­– -0.38) | -1.29$\pm$ 0.26  (-1.47– -1.11) | -- | -- | 0.07 |
| **370-470** | -1.52 $\pm$ 0.42  (-1.63 – -1.41) | -1.17 $\pm$0.43  (-1.47 – -0.87) | -1.22 $\pm$ 0.50  (-1.42 – -1.02) | -1.41 $\pm$ 0.35 (-1.64 – -1.19) | -1.41 $\pm$ 0.16  (-1.52 – -1.30 ) | **0.04*** |
| **P value** | -- | 0.08 | 0.90 | -- | -- | -- |

Values listed as mean $\pm$ SD (95% confidence interval). P values calculated with Mann-Whitney test or Kruskal-Wallis test with Dunnet’s post-hoc comparison to healthy. *: significant difference between healthy and ATP1A3 non-D801N (P=0.03). HR, heart rate; QTc, corrected QT; LOF, loss of function variant; Neg, genotype negative

**Table S13. Mean linear regression slope for HR versus QTc grouped by baseline QTc on ECG.**

| **Baseline QTc (ms)** | **Healthy** | **ATP1A3-D801N** | **ATP1A3**  **Non-D801N** | ***ATP1A3* LOF** | ***ATP1A3* Neg** | **P value** |
| --- | --- | --- | --- | --- | --- | --- |
| **<350** | -- | 1.01 $\pm$ 0.36 (0.78 – 1.23) | 0.69 $\pm$ 0.36 (0.40 – 0.98) | -- | -- | 0.07 |
| **350-370** | -- | 0.90 $\pm$0.53 (0.51 – 1.30) | 0.40 $\pm$0.27 (0.21 – 0.59) | -- | -- | 0.07 |
| **370-470** | 0.22 $\pm$0.43  (0.11 – 0.33) | 0.44 $\pm$ 0.51 (0.09 ­– 0.79) | 0.52 $\pm$ 0.53  (-0.12 – 0.07) | 0.20 $\pm$ 0.50 (0.14 – 0.49) | 0.32 $\pm$ 0.22 (0.18 – 0.50) | **0.045*** |
| **P value** | -- | 0.07 | 0.28 | -- | -- | -- |

Values listed as mean $\pm$ SD (95% confidence interval). P values calculated with Mann-Whitney test or Kruskal-Wallis test with Dunnet’s post-hoc comparison to healthy. ***:** significant difference between healthy and ATP1A3 non-D801N (P=0.01). HR, heart rate; QTc, corrected QT; LOF, loss of function variant; Neg, genotype negative

**Table S14. Characteristics of patients from IAHCRC and Baylor College of Medicine.**

|  | ***ATP1A3-*Related Phenotype*** | **ATP1A3-D801N** | **ATP1A3 Non-D801N** | ***ATP1A3* LOF**^†^ | ***ATP1A3* Neg**^‡^ | **P value** |
| --- | --- | --- | --- | --- | --- | --- |
| **Number of Individuals** | 87 | 21 | 56 | 4 | 2 | -- |
| **Number of Centers Represented** | 13 | 8 | 12 | 4 | 2 | -- |
| **Number of Holter Recordings** | 33 | 11 | 20 | 1 | 1 | -- |
| **Sex (% F)** | 54.65 | 61.90 | 53.57 | 25.00 | 100 | 0.56 |
| **Mean Age at Holter (SD)** | 17.07  (11.05) | 23.64  (9.46) | 12.87  (10.14) | 13.83 | 31.92 | 0.44 |

^*^Four patients not included in genotype groups due to unknown genotype. ^†^Includes splice site variant or deletion. ^‡^Includes no mutation identified in *ATP1A3*. P values calculated with Fisher’s exact test and Kruskal-Wallis test. LOF, loss of function variant; Neg, genotype negative.

**Table S15. Comparison of HR and QT and QTc dynamics based on flunarizine use.**

|  | ***ATP1A3*-Related Phenotypes** | **ATP1A3-D801N** | **ATP1A3 Non-D801N** | ***ATP1A3* LOF** | ***ATP1A3* Neg** |
| --- | --- | --- | --- | --- | --- |
| **% Taking Flunarizine**  **(No. Individuals)** | 68.70  (90/131)* | 73.33  (22/30) | 70.37  (57/81) | 50.00  (4/8) | 50.00  (4/8) |
| **Min HR** | 54.83, 54.20  P=0.41 | 52.61, 49.40 P=0.38 | 55.37, 56.87 P=0.99 | 59.50, 52.60 P=0.41 | 59.20, 60.00 P=0.89 |
| **Mean HR** | 92.22, 96.40  P=0.44 | 93.85, 95.20 P=0.68 | 94.39, 93.73,  P= 0.52 | 108.50, 103.80 P=0.73 | 91.60, 98.20 P=0.51 |
| **Max HR** | 158.69, 162.70  P=0.40 | 154.46, 161.80  P= 0.42 | 161.15, 155.60 P=0.30 | 159.75, 172.80 P=0.41 | 161.40, 174.80 P=0.40 |
| **HR vs QT** | -1.10, -1.29  P=0.10 | -0.88, -0.52 P=0.17 | -1.17, -1.37 P=0.26 | -1.42, -1.41 P=0.90 | -1.54, -1.30 P=0.06 |
| **HR vs QTc** | 0.63, 0.37  **P=0.02** | 0.80, 0.82 P=0.89 | 0.61, 0.33 P=0.0525 | 0.16, 0.23 P=0.90 | 0.19, 0.43 P=0.19 |

*Four patients not included in genotype groups due to unknown genotype. Mean values listed as individuals taking flunarizine, individuals not taking flunarizine, and P value using Mann-Whitney test. HR, heart rate; QTc, corrected QT; LOF, loss of function variant; Neg, genotype negative.

**Table S16. Comparison of HR and QT and QTc dynamics in patients with *ATP1A3*-related phenotypes based on flunarizine use and biological sex.**

|  | **Male**  **(flunarizine)** | **Female (flunarizine)** | ***P* value** | **Male**  **(no flunarizine)** | **Female**  **(no flunarizine)** | **P value** |
| --- | --- | --- | --- | --- | --- | --- |
| **HR vs QT** | -1.26 $\pm$ 0.45  (-1.44 – -1.08) | -0.97 $\pm$ 0.45  (-1.12 – -0.82) | **0.02** | -1.32 $\pm$ 0.44  (-1.56 – -1.07) | -1.27 $\pm$ 0.40  (-1.50 – -1.04) | 0.38 |
| **HR vs QTc** | 0.55 $\pm$ 0.44  (0.38 – 0.72) | 0.68 $\pm$ 0.56  (0.49 – 0.88) | 0.27 | 0.37 $\pm$ 0.38  (0.15 – 0.58) | 0.38 $\pm$0.48  (0.10 – 0.65) | 0.71 |

Values listed as mean $\pm$ SD (95% confidence interval). P values calculated with Mann-Whitney test. HR, heart rate; QTc, corrected QT.

**Table S17. Comparison of HR and QT and QTc dynamics in male patients with *ATP1A3*-related phenotypes based on flunarizine use.**

|  | **Male**  **(Flunarizine)** | **Male**  **(No Flunarizine)** | **P value** |
| --- | --- | --- | --- |
| **HR vs QT** | -1.26 $\pm$ 0.45  (-1.44 – -1.08) | -1.32 $\pm$ 0.44  (-1.56 – -1.07) | 0.74 |
| **HR vs QTc** | 0.55 $\pm$ 0.44  (0.38 – 0.72) | 0.37 $\pm$ 0.38  (0.15 – 0.58) | 0.13 |

Values listed as mean $\pm$ SD (95% confidence interval). P values calculated with Mann-Whitney test. HR, heart rate; QTc, corrected QT.

**Table S18. Comparison of HR and QT and QTc dynamics in female patients with *ATP1A3*-related phenotypes based on flunarizine use.**

|  | **Female (Flunarizine)** | **Female**  **(No Flunarizine)** | **P value** |
| --- | --- | --- | --- |
| **HR vs QT** | -0.97 $\pm$ 0.45  (-1.12 – -0.82) | -1.27 $\pm$ 0.40  (-1.50 – -1.04) | **0.03** |
| **HR vs QTc** | 0.68 $\pm$ 0.56  (0.49 – 0.88) | 0.38 $\pm$0.48  (0.10 – 0.65) | 0.11 |

Values listed as mean $\pm$ SD (95% confidence interval). P values calculated with Mann-Whitney test. HR, heart rate; QTc, corrected QT
